# STAKEHOLDER ENGAGEMENT IN A COMPARATIVE EFFECTIVENESS/IMPLEMENTATION STUDY TO PREVENT CA-MRSA INFECTION RECURRENCE: CA-MRSA Project (CAMP2)

**DOI:** 10.1101/2020.07.15.20147496

**Authors:** Brianna D’Orazio, Jessica Ramachandran, Chamanara Khalida, Johana Gonzalez, Rhonda G. Kost, Kimberly S. Vasquez, Teresa H. Evering, Getaw Worku Hassen, Regina Hammock, Rosalee Nguyen, Ronette Davis, Keenan Millan, Claude Parola, Barry S. Coller, Jonathan N. Tobin

## Abstract

**Background:** Methicillin-Resistant (MRSA) or Methicillin-Sensitive (MSSA) *Staphylococcus aureus* skin and soft tissue infections (SSTIs) pose serious clinical and public health challenges. Few protocols exist for outpatient education, decolonization and decontamination.

**Objectives:** This trial implemented infection prevention protocols in homes via Community Health Workers/promotoras.

**Methods:** We engaged clinicians, patients, clinical and laboratory researchers, New-York-based Federally Qualified Health Centers and community hospital Emergency Departments. The Clinician and Patient Stakeholder Advisory Committee (CPSAC) convened in-person and remotely for shared decision-making and trial oversight.

**Results:** The trial consented 186 and randomized 119 participants with SSTIs with MRSA (n=59) or MSSA (n=59), completed home visits, obtained surveillance cultures from index patients and household members and sampled household environmental surfaces at baseline and three months.

**Lessons Learned:** The retention of the CPSAC during the trial demonstrated high levels of engagement.

**Conclusions:** CPSAC was highly effective throughout design and execution by troubleshooting recruitment and home visit challenges.

## BACKGROUND

Methicillin-Resistant *Staphylococcus aureus* (MRSA) is a multi-drug resistant infection that poses serious clinical and public health challenges. As a main cause of treatment-resistant skin and soft tissue infections (SSTIs)^1,2^, MRSA carries significant morbidity and mortality, and impacts patients, families, caregivers, and health-care institutions^3,4^. While effective protocols for hospital-acquired/healthcare-associated MRSA (HA-MRSA) exist^5^, few have been adapted for use in community settings for community-associated MRSA (CA-MRSA^6–11^) which affects otherwise healthy, younger individuals without exposure to healthcare risk factors or contacts^12^. Little research has examined the feasibility and effectiveness of implementing evidence-based infection prevention interventions in primary care settings^13^ and no studies have employed Community Health Workers (CHWs) or “Promotoras” to provide home-based education and training in decolonization and decontamination. The CA-MRSA Project 2 (CAMP2) was designed to test the effectiveness and implementation of an evidence-based intervention tested and shown to be effective in the hospital intensive care unit (ICU) setting.^5^

One of the most notable features of this patient-centered outcomes research study was the engagement of multiple academic and community-based stakeholders in critical phases of the trial. The stakeholder partnership was built upon a long-term, multi-year, highly-engaged community-academic research and learning collaborative that included practicing clinicians, patients, clinical researchers, laboratory researchers, several New-York-based Federally Qualified Health Centers (FQHCs), and several Community Hospital Emergency Departments (EDs).^14^ In this article, we describe some of the logistical and procedural aspects of the community-based participatory research (CBPR) and practice-based research network (PBRN) methodologies that were used in the design and conduct of this trial and further highlight the ways in which stakeholders contributed to the CAMP2 trial.

## METHODS

The CAMP2 study was conceptualized by a group of stakeholders during a prior observational epidemiologic study, the CA-MRSA Project 1 (CAMP1), which examined the correlates, treatments, and outcomes for patients with SSTIs presenting for treatment with microbiologically confirmed *S. aureus* infections (either MRSA or MSSA).

CAMP2 was the next logical step in this CBPR partnership, where we endeavored to intervene upon patient-centered features that we observed in the CAMP1 study. The CAMP2 trial tested a community-based intervention to enhance outpatient treatment for CA-MRSA. This comparative effectiveness/patient centered outcomes research trial recruited participants at three FQHCs and three EDs in New-York City. Eligible participants: (1) were between 7-70 years; (2) were fluent in English or Spanish; (3) planned to continue to receive care in the FQHC or ED during the next year; (4) presented with signs and symptoms of a SSTI; (5) had a laboratory-confirmed baseline wound culture positive for MRSA or MSSA (patients who met inclusion criteria but whose SSTI culture was positive for *S. aureus* without antibiotic resistance, or MSSA+, were also enrolled because their risk for recurrence is similar to that of patients with MRSA+ wound cultures); (6) were willing/able to provide informed consent; and (7) willing to participate in the baseline and follow-up home visits.

Recruitment, informed consent, and baseline clinical assessment were conducted by trained CHW/Promotoras, who worked in collaboration with FQHC/ED clinicians and office staff. Data collected included clinical laboratory results from microbiological cultures taken from the participants’ wound(s), as well as surveillance cultures from the nares, axilla, and groin. Additional data collected included a detailed dermatologic assessment, clinical and demographic data extracted from the electronic health records (EHRs), molecular epidemiologic characterizations of the wound, surveillance and household specimens, environmental assessments obtained by CHW/promotoras during baseline and follow-up home visits, as well as responses to patient-reported surveys (pain, depression, quality of life, satisfaction with care).

All participants received clinician-directed standard-of-care treatment, including incision and drainage (I&D) and/or oral antibiotics. Participants were assessed at baseline and then randomized to experimental or control condition.

Two interventions were compared: (1) CDC-Guidelines directed care (i.e., incision and drainage (I&D) and antibiogram-selected oral antibiotics^15,16^ and an educational pamphlet; (Usual Care) versus (2) CDC Guidelines-directed care combined with universal household decolonization and environmental decontamination interventions based on the REDUCE MRSA Trial^5,15,16^, provided in the home by CHW/Promotoras (Experimental Group). Specifically, we evaluated the comparative effectiveness on SSTI recurrence rates (primary outcome) and secondary patient-centered outcomes (pain, depression, quality of life, care satisfaction) using a two-arm 1:1 randomized controlled trial. With the multiplicity of stakeholders’ perspectives in mind^**17**^, we arrived at the shared research priority of preventing infection recurrence and household transmission. The experimental intervention was designed to enhance participants’ MRSA knowledge and encourage self-efficacy, active self-management and preventive health behaviors^18^ (Table 1). Participants had follow-up home visits at three months and EHRs were reviewed for SSTI recurrence over six months following the index SSTI treatment.

**Table 1.** PCORI Patient-Centered Questions Addressed by CAMP2 Trial

| PCORI Patient-Centered Questions examined during CAMP1<sup>17</sup> | CAMP1 Stakeholder Feedback Addressed in CAMP2: Patient-Centered Features Incorporated into the CAMP2 Design |
| --- | --- |
| 1) <u>“Given my personal characteristics, conditions, and preferences, what should I expect will happen to me?”</u> | 78% of CAMP1 patients rated reducing the spread of MRSA in their household as “very important” to them. Moreover, 84% rated preventing their MRSA infection from coming back as “very important”. One of the goals of this project was to **reduce future recurrence** and uncertainty. Patient education and self-efficacy were crucial to the implementation of this intervention. CHW/Promotoras collaborated in developing the home visit scripts and protocol to address the cultural, socioeconomic, and medical needs of patients. |
| 2) <u>“What are my options and what are the potential benefits and harms of those options?”</u> | The CAMP2 study **compared the effectiveness of two interventions to prevent MRSA recurrence**. If effective when implemented in the community-based settings, the intervention could help reduce the spread of MRSA infection and reduce future morbidity and suffering. Given the patients who received care in the health systems settings that provide care to an urban, multi-ethnic low income population, many of whom have been disenfranchised by the health care system, all protocols were highly sensitive to participants’ autonomy and their role as the ultimate decision-maker. |
| 3) <u>“What can I do to improve the outcomes that are most important to me?”</u> | Qualitative results demonstrated that patients were most concerned about **recurrence, pain, and ability to perform functions**. CAMP2 intervention aims to empower patients to play a more active role in reducing the burden of recurrent MRSA infections through tools and methods to decolonize themselves and household members and to disinfect their households<sup>18</sup>. |
| 4) <u>“How can clinicians and the care delivery systems they work in help me make the best decisions about my health and healthcare?”</u> | Through close relationships with the communities they serve, clinicians in FQHCs and community hospital emergency departments, worked with study-supported staff, including onsite study recruiters. To minimize impact on practice workflow, research staff were present and obtained informed consent in a **collaborative style, to ensure that each participant understood that the project** was designed to help them make the best decisions for themselves, and to take active steps to reduce the possibility of infection recurrence and household transmission. |

## CBPR Methodology

### Clinician and Patient Stakeholder Advisory Committee (CPSAC) Composition and Procedures

The Clinician and Patient Stakeholder Advisory Committee (CPSAC) brought together patient stakeholders (i.e., members of the community who were not enrolled in this trial), staff from six New York City area FQHCs and EDs, staff from Clinical Directors Network (CDN, www.CDNetwork.org), a primary care practice-based research network (PBRN) and AHRQ-designated Center of Excellence for Practice-based Research and Learning (#1 P30-HS-021667), as well as academicians and clinicians from the NIH/NCATS-funded Clinical and Translational Science Award (CTSA) (#8 UL-1 TR-000043) at the Center for Clinical and Translational Science at The Rockefeller University, and scientists from the Laboratory of Microbiology and Infectious Diseases at The Rockefeller University (see Table 2). Patient stakeholders were recruited from among previous CAMP1 participants, which included participants in the observational study, focus groups and Research Town Hall meetings. In addition, the CPSAC included the designated patient representative from an FQHC, and one patient representative from each ED (n=3). CDN recruited one additional community representative to serve on the CPSAC, a local businessman and barbershop owner from previously conducted MRSA and Hepatitis C CTSA-funded pilot studies that developed and tested MRSA and Hepatitis C education with staff working at NYC barbershops^19^.

**Table 2.** Community and Patient Stakeholder Advisory Committee Roles and Composition

| Stakeholder Type | Description of Role | # FQHCs | # EDs | TOTAL |
| --- | --- | --- | --- | --- |
| Clinicians | · Conducted patient screening and recruitment<br>· Obtained informed consent<br>· Assessed dermatological symptoms, collected specimen, treated wounds, among other activities during Baseline Study Visit<br>· Assisted with follow-up of “warm handoff” protocol<br>· Participated in stakeholder meetings | 4 | 5 | 9 |
| CHW/<br>Promotoras | · Conducted patient screening and recruitment<br>· Obtained informed consent<br>· Participated in “warm handoff” protocol<br>· Conducted home visits<br>· Conducted telephone assessments<br>· Participated in stakeholder meetings | 2 | 4 | 6 |
| Patients/<br>Community Members | · Determined acceptability of education materials and home visit protocol elements<br>· Proposed community-based dissemination opportunities<br>· Reviewed assessment measures and home visit schedule<br>· Participated in stakeholder meetings | 1 | 4 | 5 |
| **TOTAL** |  | **7** | **13** | **20** |

The CPSAC met in person or via teleconference, as determined by the participants, with logistical and financial support provided by CDN. Meetings were held every 1-2 months, either in-person at The Rockefeller University (2-3 hours) or by web/teleconference (1-1.5 hours). A total of 25 CPSAC meetings were held throughout the course of the project. CPSAC roles are summarized in Table 3. During the CPSAC meetings, the team discussed study conduct and progress, identified barriers and opportunities, recommended strategies to increase recruitment, engagement, and retention of study participants, and developed opportunities for dissemination.

**Table 3.** Retention/Turnover CAMP2 Research Team

| TYPE OF STAKEHOLDER | # Beginning of study | # End of study | % Retention |
| --- | --- | --- | --- |
| FQHC/HOSP ED* | 23 | 13 | 57% |
| ACADEMIC/MEDICAL CENTER | 24 | 19 | 79% |
| PBRN | 17 | 9 | 53% |
| PATIENT/ COMMUNITY PARTNER | 5 | 5 | 100% |
| PRIVATE/CORPORATE PARTNER | 4 | 2 | 50% |
| FUNDER | 3 | 3 | 100% |
| TOTAL | 76 | 51 | 67% |

\*FQHC/HOSP ED=Federally Qualified Health Center/Hospital Emergency Department

To engage stakeholders outside CPSAC meetings, we also initiated regular communiques and encouraged stakeholders to provide ongoing input through emails and phone calls. Thus, there were numerous email threads that reflected shared decision-making. This active engagement of stakeholders fostered equitable collaboration by focusing on ongoing and multi-level communications and ensured transparency at each step. Through all stages of the project, our stakeholders shared their perspectives, preferences, and priorities.

Academic, clinician, and CHW stakeholders participated in robust discussions as to whether to conduct *S. aureus* surveillance among CHWs, based on a review of the literature and the occupational safety perspective from the CHW standpoint. Stakeholder discussions with experts in infectious diseases and infection control resulted in the decision not to conduct routine CHW surveillance, given that in settings with higher rates of MRSA exposure, such as the hospital ICU, colonization in health care workers is low^20–22^, and persistent carriage is rare^23,24^. CHW training included guidelines for infection prevention, similar to precautions taken by healthcare workers in settings with higher infection exposure and transmission risk.

## LESSONS LEARNED

### Value Added by Patient and Community Partners

The CPSAC and CHWs/promotoras routinely met to provide input and guidance on all aspects of the project. Working together, academic, clinician, CHW, and patient stakeholders made recommendations for various aspects of the trial, including but not limited to: (a) designing the home intervention, (b) selecting the primary outcomes and their measurement, (c) ensuring that patient-centered outcomes were meaningful, without being burdensome to study participants, (d) improving patient identification/recruitment, (e) obtaining informed consent, (f) intervention delivery (g) methods to improve the scheduling and completion of home visits, (g) retention goals, (h) planning dissemination activities, (i) the protocol’s burden on participants, and (j) feedback on protocol changes.

In the CAMP1 study, study participants, the research team, and clinicians identified important topics that we chose to further explore in CAMP2^14^. For example, stakeholders stressed the importance of patient education and support designed to inform participants of how the index patient and their household members could work together to implement low-cost hygienic and environmental steps to reduce the index patient’s risk of recurrent infections and prevent transmission to household members.

During the design phase of CAMP2, the stakeholders voiced strong concerns that while patients with MRSA received excellent decolonization and decontamination practices in-hospital, their needs were largely unaddressed once they left the hospital setting. At a series of community engagement meetings, stakeholders articulated their perspectives on developing a project to address CA-MRSA in the household environment, in response to a case presentation by an FQHC clinician of a CAMP1 participant with multiple recurrences.^25^ The participant was treated with I&D and antibiotics, but an SSTI recurred, and MRSA was now present in her sister who was visiting her apartment. This case study was subsequently published by one of the FQHC clinicians as the lead author^25^.

CAMP2 stakeholders and staff also discussed issues related to the conduct of home visits during early CPSAC meetings. Meeting attendees voiced concerns that participants might hesitate to invite strangers into their homes, or participants might cite “lack of trust” as a reason for failing to enroll or withdrawing from the trial. Another issue included participants’ fears of potential shaming and stigmatization about their home being “dirty” or “contaminated”. These concerns were addressed through the utilization of CHWs/Promotoras to implement the home visits, as well as in engaging the Community Health Worker Network of New York City to train study CHWs.

Stakeholders also defined the optimal process by which the project team could reach community audiences, providing input on discussions of cultural sensitivity, patient autonomy, shaming and stigmatization related to potential home contamination, and community health priorities. To address these concerns, attendees suggested having two CHWs/promotoras attend each home visit, instituting a warm hand off between clinicians and study staff, employing CHWs/promotoras who were trusted members of the community, and by explicitly addressing shaming and stigmatization in CHW/Promotora training sessions. The CPSAC also suggested additional content for training CHWs/promotoras, outlining the manner by which CHWs/promotoras should rehearse and demonstrate their competence. Academic, clinician, CHW, and patient stakeholders participated in discussions about improving patient identification and consent, methods to improve the scheduling and completion of home visits, and ways to improve the consent rate of household members.

Where approporiate, stakeholder subgroups were asked to provide input on issues germane to their expertise. CHW stakeholders tested the data collection application and refined informed consent language and assessment procedures to address language, literacy and cultural sensitivity. Community clinician stakeholders participated in refining and finalizing the study protocol, adapting and expanding the clinical workflow, and identifying patient and clinician engagement strategies. Academic stakeholders shaped discussions about patient consent and human subjects protection, the quality and acceptability of educational materials, the laboratory measures, and the patient-centered and self-reported outcomes assessment battery. They also provided input based on recently published literature on CA-MRSA and the home environment/microbiome, and guided the discussion of methods to measure intervention fidelity. Academic stakeholders also conducted ongoing discussions on building capacity for patient stakeholders to have an influence on the health of their communities. Both clinician and academic stakeholders were engaged in the development of the study-specific CHW training protocol (see Appendix D), which was implemented by an established, well-recognized CHW training organization, Community Health Worker Network of New York City (www.CHWNetwork.org). They were also involved in planning dissemination activities. Patient stakeholders were engaged in discussions about increasing the scope of dissemination venues in the community. They were additionally asked for feedback regarding protocol changes (e.g., removal of oropharyngeal surveillance swabs), as well as the acceptability of dissemination of information only (but not intervention kits), to usual care participants at the end of the study.

This active engagement of stakeholders fostered equitable collaboration through shared decision-making by focusing on ongoing and multi-level communications and ensuring transparency at each step. Our stakeholders shared their perspectives, preferences and priorities at all stages of the project. For example, in problem-solving recruitment/retention challenges, stakeholders suggested a more personalized exchange among the site clinicians, study recruitment staff and CHW/Promotoras. As such, we instituted a “warm handoff”, whereby the site clinician directly introduced the patient to the study recruiter and CHW/promotora and invited the patients to participate in the study^26^. In theory, when a patient has an established relationship with the clinician, a warm hand off by the clinician is thought to increase the likelihood that the patient will agree to participate in the study (“trust-by-proxy”). This procedure includes the patient as an active team member and engages the patient in the shared decision-making process. We found this added step to improve retention.

In addition, we were particularly eager to understand why one-third of the participants who consented to home visits withdrew from the study before they were informed about their randomized treatment assignment, and therefore never received the intervention. Home visit implementation presented a major challenge due to participants either being unreachable following their baseline visit to the FQHC or ED for treatment of their SSTIs, or they were unwilling or unable to participate due to subsequent lack of agreement by other members in the household. The perceived and actual intrusiveness of home visits proved difficult to overcome. When we shared this difficulty of retention of participants who were recruited in clinical settings but refused to participate once the project team contacted them at home, the CPSAC was instrumental in performing a “leaky pipe analysis” (see Table 4). This analysis examined the flow of patients over the study’s lifecycle, from presenting for care and informed consent to baseline home visit completion, and explored the points at which participants withdrew from the study. We undertook this analysis to improve our retention rates and to guide other community-based research projects with similar research designs. Based on stakeholder guidance we changed several procedures of the study. For example, we began making appointments at the FQHC or ED with each patient who provided informed consent (prior to laboratory confirmation), and subsequently cancelled appointments if the microbiological assessment showed that patients infection was not due to MRSA.

**Table 4.** “Leaky Pipe” Model for Evaluating Recruitment and Home Visits Completion

| Sites | # Screened | # Recruited | A<br>% Recruited of Patients Screened | # Enrolled (MRSA+/MSSA+) | B<br>% S. aureus positive | Baseline Home Visits Scheduled for All Recruited Patients^ | Baseline Home Visits Scheduled for Confirmed Eligible Patients | C<br>% Scheduled of Positive Patients | Baseline Home Visits Completed | D<br>% Home Visits Completed of # Scheduled |
| --- | --- | --- | --- | --- | --- | --- | --- | --- | --- | --- |
| FQHC A | 89 | 33 | 37% | 17 | 52% | 11 | 11 | 65% | 10 | 91% |
| FQHC B | 4 | 2 | 50% | 2 | 100% | 2 | 2 | 100% | 2 | 100% |
| FQHC C | 22 | 18 | 82% | 7 | 39% | 4 | 4 | 57% | 4 | 100% |
| ED A | 238 | 210 | 88% | 93 | 44% | 57 | 53 | 57% | 52 | 98% |
| ED B | 194 | 124 | 64% | 54 | 44% | 49 | 47 | 87% | 44 | 94% |
| ED C | 55 | 34 | 62% | 13 | 38% | 8 | 8 | 62% | 8 | 100% |
| TOTAL | 602 | 421 | 70% | 186 | 44% | 131 | 125 | 67% | 120* | 96% |
| GOAL |  |  |  | 278 |  |  |  |  | 278 |  |
| %MRSA+/MSSA+ |  |  |  | 44.1% | % of original goal (n=278) who completed baseline home visits (n=120)* |  |  |  | 43.2% |  |

^Total, including those scheduled using protocol to schedule at time of consent prior to lab result
\*One patient was determined subsequently to have HA-MRSA and was excluded from the main effects analysis (n=119)

### Continued Engagement of Patient and Community Partners

Stakeholders were highly engaged as evidenced by their enthusiasm and follow-through over the study period. We observed that the retention during the project of the CPSAC community and patient stakeholders was excellent, indicating an extremely high level of stakeholder engagement. Among the community partners, 100% remained with the project throughout the entire study and continue to collaborate as advisors in new patient-centered outcomes research studies (See Table 3). The involvement, input and continued engagement of stakeholders represent an important and integral feature of the design, conduct, and dissemination of CAMP2.

## CONCLUSIONS

CAMP2 aimed to intervene at multiple levels in the patient’s ecosystem, including the systems, patient, pathogen, and environmental factors associated with MRSA SSTI recurrence and household transmission. CAMP2 was designed based on the input of a diverse stakeholder group of practicing clinicians, patients, clinical researchers, laboratory researchers, and CHWs/promotoras. Convening the CPSAC for regular meetings gained input and guidance across all aspects of the project and encouraged sustained involvement of stakeholders in decision-making processes.

One limitation of the process evaluation of the CPSAC was the lack of a standardized tool to measure engagement and satisfaction. An engagement survey, delivered at regular intervals, could have helped quantify stakeholder satisfaction. Future analyses will examine the growth over time using network and sociometric methods.

The members of the CPSAC were instrumental from design through implementation of this comparative effectiveness/patient-centered outcomes research study. They contributed to hypothesis development and study design, as well as identified potential areas of concern during the conduct of the study, represented the patient and community-clinician points of view, and identified remedies for various study challenges (e.g., recruitment and retention). CPSAC members remained highly engaged throughout the project and were effective at strengthening the project as a whole.

## Data Availability

Data sharing is available for this trial. Contact for further details of the types of data that will be available, the available related documents, time frames, and criteria for data sharing.

## Acknowledgements

**<u>Clinical Directors Network, Inc. (CDN) Research Team</u>**

Andrea Cassells, MPH; TJ Lin, MPH; Dena Moftah, BA; Anthony Rhabb, MA, Branny F. Tavarez, MD, Cynthia Mofunanya, MD, Jasbir Singh, MBBS, MPH, Musarrat Rahman, MPH, Raul Silverio, MD, Sisle Heyliger, BA, Tameir Holder, MPH, Umamah Siddiqui, BS, Viktorya Snkhchyan MD, and Lois Lynn, MS.

**<u>The Rockefeller University Research Team</u>**

Andrea Leinberger-Jabari, MPH, Cameron Coffran, MS, Helen Marie Curry, Maija Neville Williams, MPH, Marilyn Chung, Mina Pastagia, MD, MS, Teresa L. Solomon, JD, Alexander Tomasz, PhD, Hermínia de Lencastre, PhD, and Maria Pardos de la Gandara, MD, PhD.

**<u>Federally Qualified Health Centers (FQHCs) and Hospital Emergency Departments (EDs) Community Health Network</u>**: Site Investigator: Satoko Kanahara MD, FAAP and Tyler Evans, MD.

**<u>Family Health Centers at NYU Langone:</u>** Site Investigator: William Pagano, MD, MPH, Barry Kohn MD, PhD, Isaac Dapkins, MD; Site Staff: Paula Clemons, PA; Viral Patel, MD and Jason Hyde, LMSW, MEd. Patient Stakeholder: Maria Ferrer.

**<u>Open Door Family Health Center:</u>** Site Investigator: Daren Wu, MD, Site staff: Asaf Cohen, MD. Urban Health Plan: Site Investigator: Claude Parola, MD, Tracie Urban, RN, Franco A. Barsanti, PharmD; Site Staff: Ali Saleh, PA, Scott Salvato, PA, Jennifer Concepcion.

**<u>NYC Health + Hospitals</u>**

**Coney Island Hospital ED**: Site staff: Candace Gopaul.

**Metropolitan Hospital ED:** Patient Stakeholder: Van Johnson.

**<u>Community Stakeholders:</u>**

Rosa Perez, RPh (Cordette Pharmacy) Dennis Mitchell (Denny Moe Barber Shop)

## Data and Safety Monitoring Board (DSMB)

Katherine Freeman, DrPH (DSMB Chair, Extrapolate Statistic LLC, Florida Atlantic University)

Marilyn Gaston, MD (DSMB Member, Assistant Surgeon General & HRSA Associate Administrator for Primary Health Care (ret.))

Maria Ferrer (Patient Representative)

## Scientific Consultants/Advisors

Susan Huang, MD, MPH (University of California at Irvine)

Christopher R. Frei, PharmD, MSc (STARNet/University of Texas Health Science Center at San Antonio) Eric Lofgren, PhD (Washington State University)

Christopher Mason, PhD, Ebhrahim Afshinekoo, BS, Chou Chou, MD (Weill Cornell Medicine)

Shirshendu Chatterjee, PhD, (The City University of New York (CUNY) School of Public Health)

Sarah Johnson, MD (NYC Department of Health and Mental Hygiene, Public Health and Preventive Medicine Residency Program & CUNY School of Public Health)

Bárbara Milioto, E Denise Digirolomo, Edward Clayton (Genpath/Bioreference Labs) Vicky Seyfert-Margolies, PhD, Dana Wershiner, Trang Gisler, MS (MyOwnMed, Inc.)

Suzanne Lechner, PhD, Sara Vargas, PhD, Suzanne Hower, PhD (Consulting Associates, Inc.)

## Funding Agency

**<u>Patient Centered Outcomes Research Institute (PCORI):</u>** Anne Trontell, MD, MPH, Donna Gentry, MA and Jess Robb, MPH.

## REFERENCES

1. Moran GJ, Krishnadasan A, Gorwitz RJ, et al. Methicillin-resistant S. aureus infections among patients in the emergency department. N Engl J Med. 2006;355(7):666–674.

2. Farr AM, Aden B, Weiss D, Nash D, Marx MA. Trends in hospitalization for community-associated methicillin-resistant Staphylococcus aureus in New York City, 1997-2006: data from New York State’s Statewide Planning and Research Cooperative System. Infect Control Hosp Epidemiol. 2012;33(7):725–731.

3. Klevens RM, Morrison MA, Nadle J, et al. Invasive methicillin-resistant Staphylococcus aureus infections in the United States. Jama. 2007;298(15):1763–1771.

4. Centers for Disease Control and Prevention. Methicillin-resistant Staphylococcus aureus (MRSA) infection in healthcare settings. http://www.cdc.gov/HAI/organisms/mrsa-infection.html. Published 2012. Accessed February 2, 2019.

5. Huang SS, Septimus E, Kleinman K, et al. Targeted versus universal decolonization to prevent ICU infection. N Engl J Med. 2013;368(24):2255–2265.

6. Tenover FC, Goering RV. Methicillin-resistant Staphylococcus aureus strain USA300: origin and epidemiology. J Antimicrob Chemother. 2009;64(3):441–446.

7. Cluzet VC, Gerber JS, Metlay JP, et al. The Effect of Total Household Decolonization on Clearance of Colonization With Methicillin-Resistant Staphylococcus aureus. Infect Control Hosp Epidemiol. 2016;37(10):1226–1233.

8. Huang SS, Septimus E, Kleinman K, et al. Chlorhexidine versus routine bathing to prevent multidrug-resistant organisms and all-cause bloodstream infections in general medical and surgical units (ABATE Infection trial): a cluster-randomised trial. Lancet (London, England). 2019;393(10177):1205–1215.

9. Papastefan ST, Buonpane C, Ares G, Benyamen B, Helenowski I, Hunter CJ. Impact of decolonization protocols and recurrence in pediatric MRSA skin and soft-tissue infections. J Surg Res. 2019;242:70–77.

10. Fritz SA, Hogan PG, Hayek G, et al. Household versus individual approaches to eradication of community-associated Staphylococcus aureus in children: a randomized trial. Clin Infect Dis. 2012;54(6):743–751.

11. Ellis MW, Griffith ME, Dooley DP, et al. Targeted intranasal mupirocin to prevent colonization and infection by community-associated methicillin-resistant Staphylococcus aureus strains in soldiers: a cluster randomized controlled trial. Antimicrobial agents and chemotherapy. 2007;51(10):3591–3598.

12. DeLeo FR, Otto M, Kreiswirth BN, Chambers HF. Community-associated meticillin-resistant Staphylococcus aureus. Lancet. 2010;375(9725):1557–1568.

13. Fritz SA, Camins BC, Eisenstein KA, et al. Effectiveness of measures to eradicate Staphylococcus aureus carriage in patients with community-associated skin and soft-tissue infections: a randomized trial. Infect Control Hosp Epidemiol. 2011;32(9):872–880.

14. Kost RG, Leinberger-Jabari A, Evering TH, et al. Helping basic scientists engage with community partners to enrich and accelerate translational research. Academic medicine : journal of the Association of American Medical Colleges. 2017;92(3):374–379.

15. Stevens DL, Bisno AL, Chambers HF, et al. Practice guidelines for the diagnosis and management of skin and soft tissue infections: 2014 update by the Infectious Diseases Society of America. Clin Infect Dis. 2014;59(2):e10–52.

16. Liu C, Bayer A, Cosgrove SE, et al. Clinical practice guidelines by the Infectious Disease Society of America for the treatment of methicillin-resistant Staphylococcus aureus infections in adults and children. Clin Infect Dis. 2011;52(3):e18–55.

17. Patient-Centered Outcomes Research Institute. Patient-Centered Outcomes Research. https://www.pcori.org/research-results/patient-centered-outcomes-research. Published 2013. Accessed 2 February 2019.

18. Pardos de la Gandara M Raygoza Garay JA, Mwangi M, et al. Molecular types of methicillin-resistant Staphylococcus aureus and methicillin-sensitive Staphylococcus aureus strains causing skin and soft tissue infections and nasal colonization, identified in community health centers in New York City. J Clin Microbiol. 2015;53(8):2648–2658.

19. Leinberger-Jabari A, Kost RG, D’Orazio B, et al. From the bench to the barbershop: community engagement to raise awareness about community-acquired methicillin-resistant Staphylococcus aureus and Hepatitis C virus infection. Progress in community health partnerships : research, education, and action. 2016;10(3):413–423.

20. Dulon M, Peters C, Schablon A, Nienhaus A. MRSA carriage among healthcare workers in non-outbreak settings in Europe and the United States: a systematic review. BMC infectious diseases. 2014;14:363.

21. Johnston CP, Stokes AK, Ross T, et al. Staphylococcus aureus colonization among healthcare workers at a tertiary care hospital. Infect Control Hosp Epidemiol. 2007;28(12):1404–1407.

22. Price JR, Cole K, Bexley A, et al. Transmission of Staphylococcus aureus between health-care workers, the environment, and patients in an intensive care unit: a longitudinal cohort study based on whole-genome sequencing. Lancet Infect Dis. 2017;17(2):207–214.

23. Cookson B, Peters B, Webster M, Phillips I, Rahman M, Noble W. Staff carriage of epidemic methicillin-resistant Staphylococcus aureus. J Clin Microbiol. 1989;27(7):1471–1476.

24. Siegel JD, Rhinehart E, Jackson M, Chiarello L, Healthcare Infection Control Practices Advisory C. Management of multidrug-resistant organisms in health care settings, 2006. Am J Infect Control. 2007;35(10 Suppl 2):S165–193.

25. Balachandra S, Pardos de la Gandara M, Salvato S, et al. Recurrent furunculosis caused by a community-acquired Staphylococcus aureus strain belonging to the USA300 clone. Microb Drug Resist. 2015;21(2):237–243.

26. Agency for Healthcare Research and Quality. Warm handoff: intervention. U.S. Department of Health and Human Services. https://www.ahrq.gov/professionals/quality-patient-safety/patient-family-engagement/pfeprimarycare/interventions/warmhandoff.html. Published 2017. Accessed January 15, 2017.

